# An Environmental Scan of American and Canadian Translational Science Training Programs at the Graduate Level

**DOI:** 10.1101/2023.04.09.23288339

**Authors:** Charlotte Ouimet, Adam S. Hassan, Christina Popescu, Vivienne Tam

**Author notes:** **Corresponding Author:** Name: Vivienne Tam, Address: 2250 Wong Building, 3610 Rue University, Montreal QC H3A0C5.

## Abstract

To date, there are considerable delays in bringing academic innovations into clinical practice. In part, this is due to a lack of knowledge translation and communication between clinicians and scientists. While MD/PhD programs could bridge this gap, more inclusive and sustainable alternatives must be explored. In the United States, the Howard Hughes Medical Institute (HHMI) launched an initiative to create programs wherein graduate students would be exposed to clinical curricula and establish networks with health professionals. In this study, we aim to survey such programs in North America and identify key features. In our environmental scan, we analyzed the translational science training curricula of 28 American and 17 Canadian universities. We observed that 25 schools in the United States offered training in translational science at various degree levels (certificate, Master’s, PhD, etc.) whereas only 4 Canadian institutions did so and primarily at a Master’s level. From those programs, 5 American universities offered a multi-faceted training program that met at the intersection of courses, clinical mentorship, and networking opportunities compared to only 1 in Canada. Therefore, while we noted a growing interest in science translation programs in the United States, there is a current lack of such programs at Canadian institutions. Based on the need established by this environmental scan, we hope to establish a translational science certificate program at McGill University that fills this training void and paves the way for other universities across Canada.

## Introduction

Despite biomedical research advancing at a breakneck pace, we have often failed to effectively translate these findings into tangible improvements in human health (MacLeod, 2014). Work has shown that it takes an average of 17 years for research evidence to be included in clinical practice (Comeau et al., 2017). This means that despite the resources and time invested in basic research, it takes close to two decades for any advance in basic research to impact the care we provide patients, illustrating the urgent need for better knowledge translation from the basic sciences into clinical practice. The primary barriers to effective knowledge translation include (1) a lack of scientists able to effectively translate their research and (2) the lack of infrastructure required to facilitate interdisciplinary communication, particularly between clinicians and scientists (Heller & De Melo-Martín, 2009). Over time, MD/PhD programs have become the most visible training options for such interdisciplinary training (Brass et al., 2010). However, given how time-intensive, resource costly and selective these programs are, this approach alone does not meet the growing need for effective translational science at the institutional level (Smith et al., 2013).

Alternatively, we need to focus on translational science training programs as a means of increasing knowledge translation. Current specific translational training and career incentive in the field is lacking (Jane Budge et al., 2015). Thus, a rethinking of how we train the next generation of translational scientists is imperative if we are to break down knowledge silos between disciplines and facilitate the flow of innovation from bench to bedside. Not only would a new translational science training model ensure that basic science discoveries reach the care of patients, but it would allow physicians to be involved in scientific discovery without being basic scientists. Basic scientists would, in turn, have a better understanding of the needs of patients and how best to curate their research to lead to impactful change. Previous programs have demonstrated that clinicians who receive extra support from their institutions to mentor graduate students have a better rapport with them which increases the flow of information between basic science and clinical science (Smith et al., 2013). An increased focus on translational science training would foster the collaborative relationships needed so that every stakeholder in medicine had the opportunity to contribute to the scientific dialogue.

Such translational training programs have received funding in the United States. In an initiative supported by the non-profit Howard Hughes Medical Institute (HHMI), 23 programs were created at US-based institutions to expose graduate students to medicine, and provide them with the language, culture, and network of the clinical world. This grant awarded institutions $16 million USD ($700,000 USD over 4 years) to “integrate medical knowledge and an understanding of clinical practice into their biomedical PhD curricula” (*The 2010 Med into Grad Grantees* | *HHMI*, n.d.). The aim of these programs was to reduce the research-to-practice gap by empowering students to engage in knowledge translation efforts through exposure and networking opportunities between emerging scientists and clinicians.

Despite the existence of funding sources for potential translational science training programs, to our knowledge, little work has been performed to survey the North American landscape of effective longitudinal educational programming for graduate science students designed to support knowledge translation. While several piecemeal opportunities are often available in science graduate programs, a longitudinal program places the focus on training scientists to understand the world of medicine and how they can meaningfully engage with clinicians and the clinic through networking opportunities, guided peer discussion on clinical observations, and tailored interactive coursework.

As the first step in our own educational design efforts for a translational certificate program at McGill University, we engaged in an environmental scan and benchmarking exercise. The aim of our study was to identify key characteristics of North American longitudinal knowledge translation programs available to science graduate students to (1) determine the current landscape of translational science training programs in Canada and (2) identify strong features of existing programs that should be considered in the creation of our own program. With a focus on longitudinal programs, we identified and analyzed available programs at a variety of schools according to their degree level, duration, and content. By synthesizing available programs, we provide a benchmarking foundation for others who wish to implement similar programs at their institutions.

## Environmental scan

Given our target program will be designed for a Canadian student audience, our primary objective was to document the existence and nature of translational science programming currently available in Canada. We quickly realized we had to widen our scope as a means of better understanding the greater context of the interplay between basic science graduate programs and clinical programs and knowledge. We therefore turned our attention to a subset of the institutions in the United States, given the vast similarities in university infrastructure but more extensive programming in the U.S. due to the HHMI grant. We began by identifying universities of interest. The American universities selected in our cohort consisted of those that received the HHMI “Med into Grad” grant in 2010; a grant awarded to schools to help develop new graduate programs or enhance existing programs to train translational scientists^7^ (n=28). The schools that received this grant had both an interest and an intention to create academic programs aimed at educating translational scientists. We complemented our search with 5 additional American universities that our institution commonly includes in similar benchmarking exercises since they are all Association of American Universities (AAU) public institutions with similar profiles (size, funding, etc.), or represent aspirational institutions with more resources. We did not perform an exhaustive analysis of all American universities since the primary aim of our study was to analyse the translational education landscape in Canada. Therefore, the American arm of our research serves to complement the analysis we performed of Canadian universities. As for the Canadian universities selected, we chose to analyse all universities with accompanying medical schools (n=17) given that they have the necessary knowledge and resources to support translational science programming.

## Methodology

For all the universities in our cohort, we performed a web search in January 2021 using Google as a search engine with the following search terms: “Translational Science Certificate Program” OR “Med into Grad” OR “Translational Research” OR “Translational Training” OR “Translational Medicine” OR “Translational Science” OR “Multidisciplinary Training” AND university name. For each university, we identified the translational program offered and collected the following information: (a) name of the program, (b) length of the program, (c) nature of the program (Certificate, Master’s, Ph.D., Other), (d) duration of the program, (e) target population for the program, (f) if the program incorporated clinical mentorship, courses and/or networking opportunities. When multiple translational programs were offered by one university, we selected the translational education program that provided the most holistic training opportunities for graduate science, technology, engineering, and math (STEM) students. Data analysis was double coded by two individuals (CO, ASH). The data were consolidated and all disagreements were resolved by discussion and consensus.

## Results

Twenty-eight American schools and seventeen Canadian universities were included in our analysis. Ninety-three percent (n=26) of American schools examined and 23.5% (n=4) of Canadian schools had a longitudinal translational program. Of the 26 institutions, one program was shared by two institutions (University of California Berkeley and University of California San Francisco), therefore, a total of 25 programs are captured in the analysis.

Of the 25 American and 4 Canadian programs in translational research, we assessed whether matriculants were granted certificates, Master’s, or doctoral degrees or others upon completion. In Canada, all 4 universities offer programs at the Master’s level. Of the four, one University offers its program at the Ph.D. level as well. Ten American universities offer certificate programs (40%), 4 provide Master’s programs (16%), 3 provide doctoral programs, and 8 were unspecified and termed as ‘other’ (32%) (Figure 1A). Of those 8, most are to be completed in conjunction with an advanced degree (e.g. Ph.D. in biomedical sciences) or offered to post-doctoral fellows or physicians.

**Figure 1.**
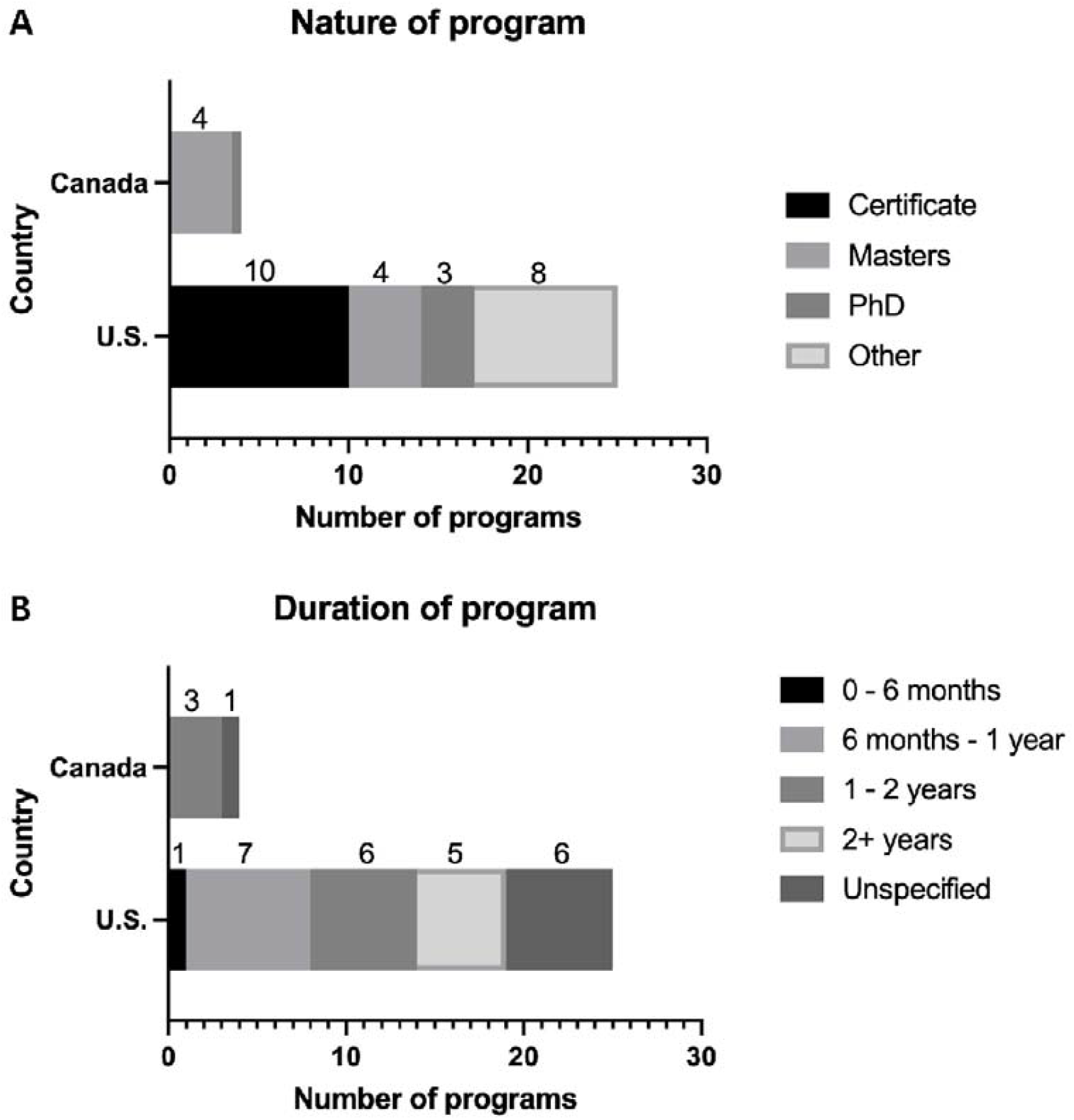
Characteristics of Translational Research Programs in North America. Nature (A) and duration (B) of programs per country of origin.

To assess the duration of translational programs, we divided the programs into 5 categories: 0 – 6 months, 6 months – 1 year, 1 – 2 years, 2+ years, and ‘unspecified’ (Figure 1B). Programs lasting an indefinite amount of time were typically the result of a certificate program or ‘other’ types of curricula that were offered alongside a doctoral program and considered completed at the end of a student’s graduate studies.

In addition to differing in degree level and duration, we observed a disparity in how the program structures incorporated courses & didactic instruction, networking opportunities, and/or clinical mentorship. We assessed the number of schools that provided either a single axis or a multidimensional approach. For Canadian schools, 2 programs were strictly course-based (50%) whereas 1 combined courses and networking (25%), and the last one combined all three axes. Meanwhile, 12 American programs were primarily course-based (48%), 2 emphasized solely clinical mentorship (8%), and 1 was focused on networking (4%). In addition, 5 programs from American universities combined coursework and clinical mentorship (20%) whereas 5 institutions provided instruction combining the three main axes in some form (20%) (Figure 2).

**Figure 2.**
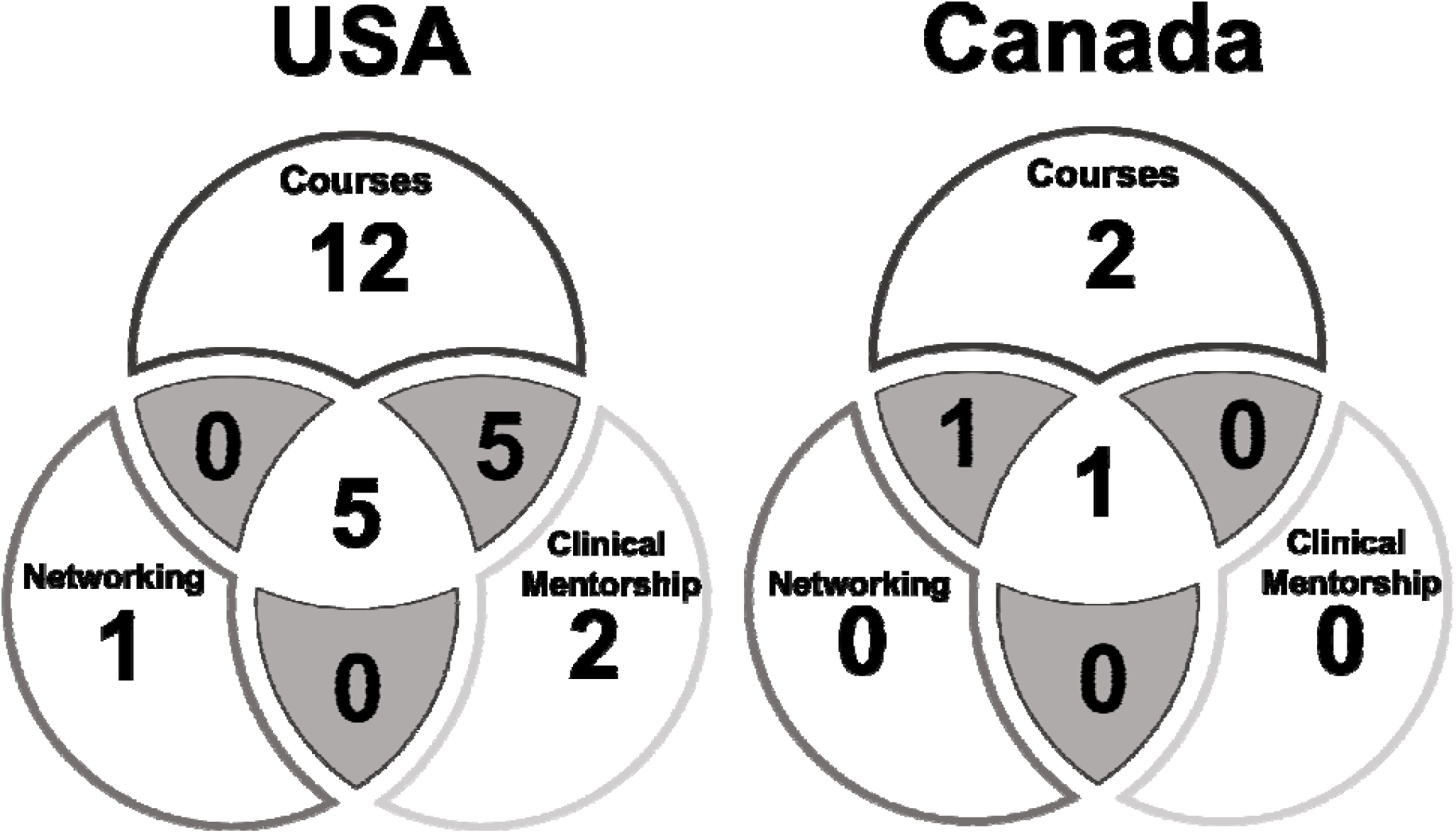
Translational Research Program Content. Comparing the number of programs that are course-based, networking-based, and clinical mentorship-based programs in the United States (left) and Canada (right).

## Local context

The long-term goal is to create a longitudinal program at our institution - McGill University. Currently, students interested in translational research can take individual courses or attend workshops, but the offerings are sparse and department-specific, and thus not widely available to the STEM student body. We would like to create a translational certificate program at our institution to fill this void. During our benchmarking process, we identified the Leder Human Biology and Translational Medicine program run out of Harvard University as being the closest example of the certificate program we aim to create and, thus, the model for our novel program (*Admissions - by the Numbers* | *Office of Admissions - McGill University*, n.d.). It is a 1.5-year certificate program that doctoral students can opt into in their first year of graduate studies through department affiliation with the Harvard Integrated Life Sciences Federation. In the first three semesters, students engage in pre-clinical coursework specifically crafted for the biomedical scientist in mind, while still aiming to offer students a solid foundation in human physiology and pathology. In the remainder of the program, students are paired with physicians to learn the culture of medicine and the roles basic scientists can have in the clinical setting.

## Discussion

The overarching goal of this investigation was to equip us with evidence-based knowledge needed to create a translational certificate program for graduate STEM students at McGill University. To that end, we aimed to analyze programs from similar universities in the U.S. and Canada to gain a better understanding of various opportunities open to aspiring translational scientists, who wish to enhance their training with clinical knowledge as a means of guiding their research and improving knowledge translation.

Our research showed that (1) there are few schools offering a translational program in Canada (n=4) and, (2) programs in the U.S. offer various formats ranging from Master’s or PhD programs to certificates completed concurrently with the student’s existing degree, (3) the Canadian programs are mostly 1-2 year Master’s programs (75%) (Figure 1A). Notably, there is no option in Canada for students to receive institutional training in translational research while completing a separate graduate degree.

As demonstrated by the uptake of the translational programs in the United States, there is a current interest for translational training programs. Out of the initial 23 American institutions awarded the HHMI program in 2010, 21 still have at least one translational program for graduate STEM students, their longevity illustrating the interest and success of such programs once they have been established. In Canada, there is currently a lack of such training programs. Out of the 17 medical schools in Canada, translational programs are only offered in 4 of those, most of which are masters programs (Figure 1A). Anyone aiming to pursue translational science needs to commit to a full-time program which cannot be done concurrently with other graduate training if they aim to receive recognition for their training. As opposed to fostering collaborations between disciplines, these programs continue to endorse this idea that you must enter a specific silo to practice translational science. We argue that a framework needs to exist in which any interested scientist can obtain the skills required to integrate their work into a broader knowledge framework without needing to pause their research.

Another option for aspiring translational scientists is the MD/PhD program offered by many universities in both the United States and Canada. However, these programs are not a sufficient training method to satisfy the growing need for translational scientists. Not only has the Canadian Institutes of Health Research (CIHR) terminated funding for the programs in 2016 (Lewinson et al., 2016), but the remaining programs are extremely small and selective. For example, McGill University only admitted 5 students in 2019/2020 in their MD/PhD program (McGill University). In addition, these programs are large commitments with the average time of completion of 8.0 years (Brass et al., 2010; Brass & Drubin, 2018). Limited funding for an already small program means that it cannot respond to the need for translational science training. Thus, if Canada is to be competitive in the field of translational science, it is forced to re-examine how it is broadening the accessibility of translational science training to equip more basic scientists, as well as deepening the scope of what is being offered in these programs.

As translational scientists build relationships that bridge knowledge silos and learn the language of medicine to effectively engage and communicate with the clinical world, the clinical mentorship and networking activities become increasingly important components of more holistic, translational training. Canadian programs are primarily oriented around courses. A successful translational program requires the integration of three essential aspects: course-based, networking, and clinical mentorship, and very few programs we surveyed (see Figure 2) exist at this intersection, with a large majority being solely course-based (n = 14). If we look closer at the programs that successfully integrate all three aspects (see full list in Supplementary Table 1), 5 are located in the USA and 1 in Canada. Thus, in the educational design of the McGill program, we aim to integrate all three essential aspects found in the programs we surveyed.

## Conclusion

In this study, we benchmarked 17 Canadian universities and found that four (23.5%) offered any type of translational program. All four universities offered a Master’s program with one university also offering a PhD program. This differed from American schools which offered an array of certificate programs, Master’s programs, doctoral training programs with 5 of those universities incorporating clinical mentorship, networking in addition to coursework. Only 1 of the Canadian schools offered a program that included those three pillars. Most notably, no translational training program exists in Canada that can be done concurrently with another graduate program. Our research has therefore demonstrated that there is a gap in the translational science education offered in Canada, which McGill University has an opportunity to fill with the development of a translational science certificate program. We envision and aim to implement a 1.5-year program that will enrich basic science training through a mix of medical-style coursework crafted for graduate students, an immersive clinical experience and engagement with the broader translational network at McGill. Translational programs like the one we hope to create are a pivotal part of training a generation of scientists who know how to bridge silos of knowledge, as well as building infrastructure that can support effective translation. We hope this analysis has helped to illustrate and better define this pressing pedagogical need in Canadian universities. As we develop this program, we will continue to share our pedagogical method and findings, in the hope of sparking impetus and know-how amongst students who wish to do the same in their own universities. It is not often that students get to take ownership of their own education in this way, but we believe this work will help to empower students globally to build more effective translational communities in their local context.

## Data Availability

All data produced in the present study are available upon reasonable request to the authors.

## Acknowledgments

The authors would like to thank Dr. Meredith Young for her insightful comments on the manuscript. They would also like to thank Dr. Terry Hébert and Ms. Leigh Dickson for their faculty leadership and guidance in developing this program.

## Supplementary Information

**Supplementary Table 1.**
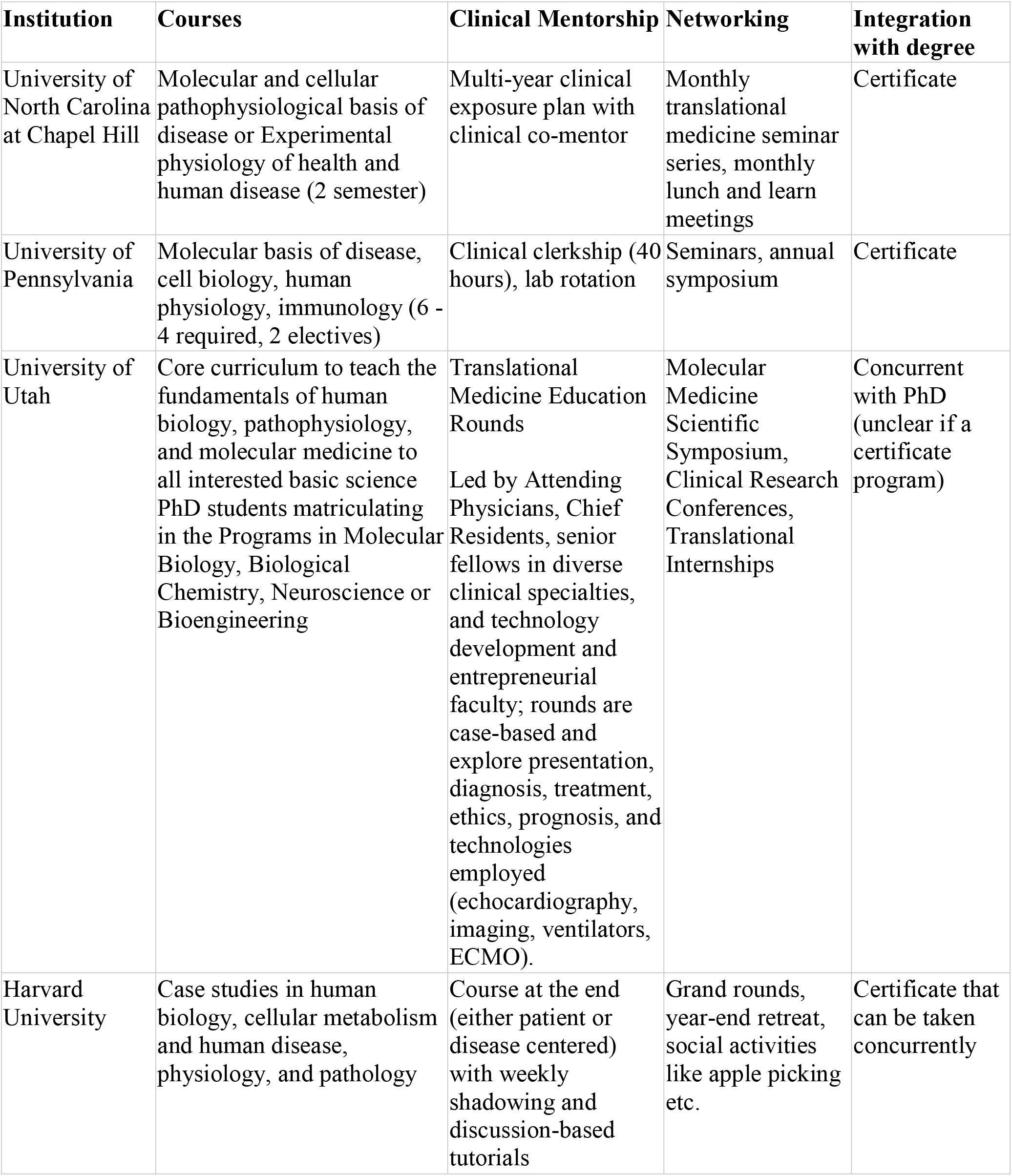
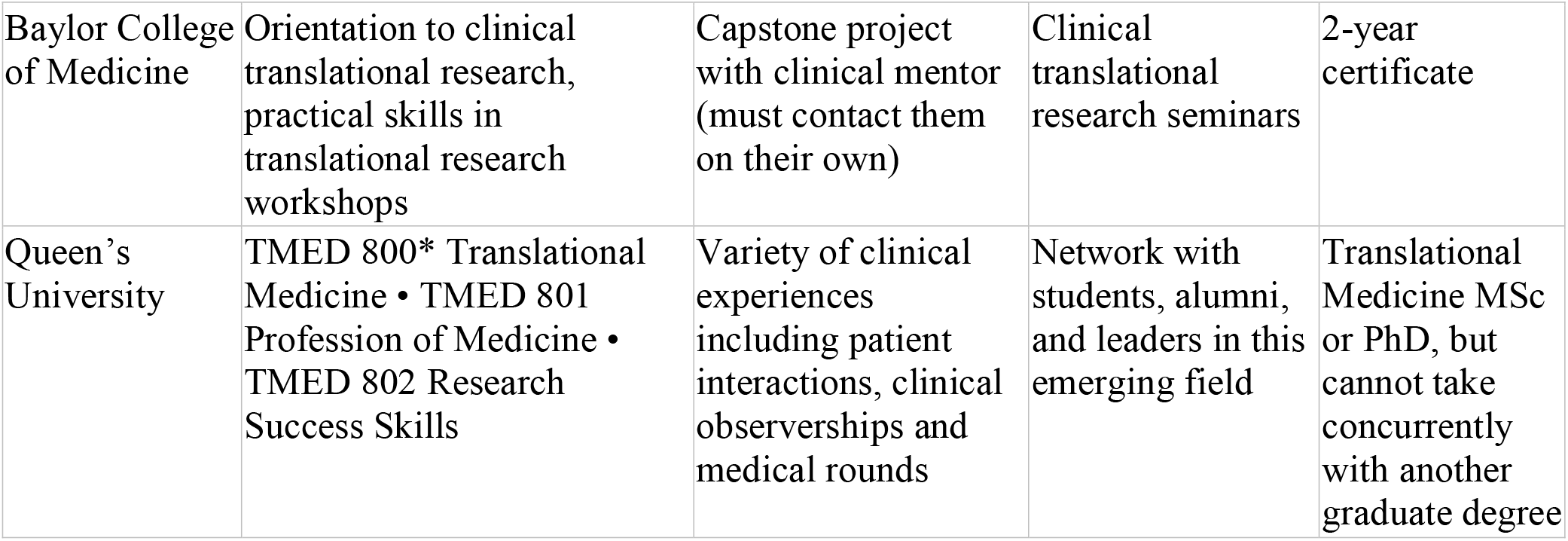
Additional information regarding schools with translational science programs incorporating all three factors. 5 American universities and one Canadian institution from Figure 2 offer programs that lie at the intersection of course-based learning, networking, and clinical mentorship.

